# A Pause, Not a Stop: Language Regression in Toddlers at High Familial Likelihood of Autism

**DOI:** 10.64898/2025.12.08.25341837

**Authors:** Margaret L. McAllister, Tyler McFayden, Shruthi Ravi, Lonnie Zwaigenbaum, Robert Schultz, Annette Estes, Jessica Girault, Mark Shen, Meghan R. Swanson

**Affiliations:** University of North Carolina at Chapel Hill, Department of Health Sciences, Division of Speech and Hearing Sciences, Chapel Hill, NC, USA; University of North Carolina at Chapel Hill, Carolina Institute for Developmental Disabilities, Chapel Hill, NC, USA; University of Washington, Institute on Human Development and Disability, Autism Center, Seattle, WA, USA; Department of Pediatrics, University of Alberta, Edmonton, AB, Canada; Center for Autism Research, Children’s Hospital of Philadelphia, University of Pennsylvania Perelman School of Medicine, Philadelphia, PA, USA; University of Washington, Department of Speech and Hearing Sciences, Seattle, WA, USA; University of Minnesota, Department of Pediatrics, Twin Cities, MN, USA

**Author notes:** Correspondence to: Meghan Swanson, PhD 2025 E. River Parkway 7962A, Minneapolis, MN 55414.

## Abstract

Language development, a core pillar of social communication, has variable trajectories in autism that include a regression or loss of skills in roughly 20% of autistic individuals. Language regression is most frequently identified through parent report but can also be observed as a decrease in raw scores on a repeated language assessment (measure-defined). Later language outcomes after regression have been observed to be highly variable, but not lower than children without a language regression. The current study explores rates of parent-reported and measure-defined language regression in a large sample of infants at high familial likelihood of autism due to having an older autistic sibling. Among all participants at high familial likelihood for autism (*n*=428), parent-reported regression was observed in 2.8% (*n*=12) and was associated with 2.77 times higher odds of receiving an autism diagnosis. Measure-defined regression was observed in 8% (*n*=36) and was associated with 1.21 times higher odds of autism diagnosis. These rates of regression are expectedly lower than estimates collected in autistic samples. Neither of these elevated odds was statistically significant and there was low concordance between these groups with only one participant present in both. Nearest-neighbor comparison samples of non-autistic infants at high and low likelihood for autism without language regression were selected to assess differences in language growth trajectories associated with regression. Infants with parent-reported language regression showed comparable language development to a matched high-likelihood sample while infants with measure-defined language regression showed slower overall language development than matched peers. Taken together, our results show that parent-report and direct measurement of regression capture unique aspects of child language development that may not be predictive of an autism diagnosis but may indicate delayed language growth in early toddlerhood. These language outcomes support previous findings of wide heterogeneity among those with regression and continued language growth after loss of skills.

**Key Points:**

- Language regression can be captured through parent-report or decrease in raw scores on repeated language assessment and is reported in approximately 20% of autistic toddlers.
- Most research on language regression uses retrospective report of regression in autistic children, but this study prospectively examines regression in toddlers at high familial likelihood for autism who do and do not receive later diagnoses.
- Parent-reported and measure-defined regression in this high-likelihood sample have low concordance indicating that these may be different events in language development.
- The presence of language regression was not associated with significantly higher odds of receiving an autism diagnosis.
- Children who exhibit language regression continue growing and developing language and those with parent-reported regression display comparable language skills to children without language regression at three years of age.

## Introduction

Autism spectrum disorder (hereafter, *autism*) involves differences in social communication and repetitive behaviors (APA, 2013). Language, a core pillar of social communication, is highly variable in autism, with the language ability of autistic individuals’ ranging from no expressive language to highly proficient and above average verbal skills (Elsabbagh, 2020). Autistic individuals also demonstrate highly variable trajectories of language development, such that some with delays in language catch up in later childhood (Pickles et al., 2022). Because of this heterogenous developmental landscape in autism, language represents a domain of high interest for both research and clinical communities. Delays in language are often the first core concern reported by parents (Becerra-Culqui et al., 2018) and attaining phrase speech is one of the best predictors of later quality of life in autistic individuals (Mayo et al., 2013), which underscores the interest and importance of understanding language trajectories in autism.

One linguistic phenotype associated with autism is regression, defined as the loss of previously acquired words or phrases (Barger et al., 2013; Ozonoff & Iosif, 2019). Language regressions are not unique to autism and can often be seen in other disorders, such as epileptic aphasia (Landau-Kleffner Syndrome; Deonna & Roulet-Perez, 2010; Shinnar et al., 2001). However, language regressions do appear to have unique phenotypes in autistic children (Pickles et al., 2022) that do not appear in other disorders such as developmental language delays (Pickles et al., 2009). Language regression is a subset of skill losses that have been observed in autism, including regressions in socialization, fine/gross motor, or eating/toileting behaviors (Barger et al., 2013). In a meta-analysis spanning 29,000 autistic participants, rates of regression varied by sampling method and domain. The overall prevalence rate for regressions ranged from 21.8%-40.8% and prevalence of language regression specifically was estimated at 24.9% (Barger et al., 2013). A meta-analytic update from Tan and colleagues (2021) provided similar estimates that approximately 30% of autistic youth experience a regression when pooling across all domains; when looking at language specifically, the prevalence rate is approximately 20%.

Individuals who experience a language regression tend to walk earlier and use first words earlier but are not significantly different than non-regression groups on autism severity or presentation of autism-specific traits (Jones & Campbell, 2010; Manelis et al., 2020; Meilleur & Fombonne, 2009; Pickles et al., 2022). Considering later outcomes, individuals who experience language regressions often attain phrase speech at comparable times to those without regressions (Pickles et al., 2022; Prescott & Weismer, 2021), although there is significant variability. When following language-regressed youth to age 10 years in a prospective longitudinal study, Pickles and colleagues (2022) concluded that although there was considerable variability in language outcomes, many youth developed average to above-average outcomes compared to autistic youth without previous language regression. Thus, although language regressions are reportedly concerning for parents (Clarke, 2019), research suggests that early language regression does not significantly negatively impact subsequent language development. However, language regressions may be signals to providers and parents that additional intervention or speech-language therapy would be beneficial to attenuate the regression’s potential impacts on later growth. As such, recognizing and detecting language regression is the first step towards swift action.

Language regression is most commonly measured using a parent-report form called the Autism Diagnostic Interview-Revised (ADI-R; Rutter et al., 2003), which provides a systematic framework for recording parent-reported regressions after the first 3-5-word stage of acquisition (Tan et al., 2021). Results from the meta-analytic update (Tan et al., 2021) indicated that language regression is largely assessed using the ADI-R. Other methods include other types of parent interview, review of medical records, and observation through direct assessment, wherein regression is defined by any raw score decrease in language scores between two timepoints (R. Landa, 2007; R. Landa & Garrett-Mayer, 2006; R. J. Landa et al., 2013; Ozonoff & Iosif, 2019). Parent-report and direct assessment have not been compared and therefore it is possible that they may capture different behaviors (Tan et al., 2021). Direct assessment is subject to some measurement error due to child factors like fatigue, hunger, illness, and mood. Meanwhile, parent-report is subject to varied interpretations of what counts as a regression. For example, a lack of growth or stagnation could be interpreted as a “regression” (Clarke, 2019). Parents may also have a retrospective bias when looking back several years (Goldberg et al., 2003) if asked about regression during childhood.

The field largely lacks a standardized definition and direct measure for language regression, which was one of the conclusions from Tan et al., (2021), calling for the “development of a standardized tool… to measure language regression at multiple time points during early childhood development.” (Tan et al., p. 592). The current longitudinal study characterizes early childhood language trajectories prospectively from 6-36+ months in a sample enriched for autism by using an infant sibling research design. This approach to studying language regressions is novel given that most of the work to date has involved retrospective reporting. When prospective studies have been used, the youngest age group has been 2-5 years (Pickles et al., 2022), which is after the average age of language regression onset. The current study uses measurements of parent-report (ADI-R) and repeated, objective measures of language between 3 and 36 months of age (Mullen Scales of Early Learning; Mullen, 1995) to evaluate whether standardized behavioral assessments are sensitive to parent-reported language regressions during this developmental period. Furthermore, we used these measures to determine whether language regressions were present in toddlers whose parents do not report a regression. Using a parent-reported *and* measure-defined approach to investigate language regressions in infants with and without later autism diagnoses will provide valuable insight into the early developmental trajectories of receptive and expressive language.

The aims of the current study were to explore the phenotypes of language regressions in children at high familial likelihood of autism (indicated by having an older, autistic sibling):

1. Evaluate the frequency of language regression and concordance between parent-reported and measure-defined regressions;
2. Investigate the diagnostic outcome of toddlers with parent-reported and measure-defined language regression and assess differences in language outcomes by diagnostic group; and
3. Characterize longitudinal trajectories of language ability in infants with language regressions compared to nearest-neighbor matched groups without language regression to evaluate significant differences in language growth and outcomes over the first three years of life.

## Method

### Data Source

Participant data was collected through a prospective, longitudinal study of infants at high and low likelihood for autism called the Infant Brain Imaging Study (IBIS). Autism likelihood status was defined by having an older sibling with a diagnosis of autism (HL) or an older typically developing sibling (LL). The study was conducted at four sites: University of North Carolina at Chapel Hill, Children’s Hospital of Philadelphia, University of Washington, and Washington University with data coordination at Montréal Neurological. All study procedures were approved by the institutional review board at each collection site and written informed consent was obtained from the guardians of all participants.

Exclusion criteria for the IBIS study were (1) evidence of a genetic condition, (2) vision or hearing impairment, (3) gestational age < 36 weeks and/or birth weight < 2000 g, (4) significant perinatal adversity or prenatal toxin exposure, (5) contraindication for magnetic resonance imaging (MRI), (6) primary home language other than English, (7) sibling statuses of adopted, half-sibling, or twins, and (8) first-degree relatives with diagnoses of psychosis, schizophrenia, or bipolar disorder. To be eligible for inclusion in the current study, participants in the HL group were required to (a) be at high familial likelihood for autism, (b) have completed an autism evaluation, (c) have completed the Mullen Scales of Early Learning (MSEL; Mullen, 1995) at two or more timepoints, and (d) have completed the ADI-R at 24 and/or 36 months of age. Low likelihood participants considered for inclusion as a matched sample (a) completed the MSEL at two or more timepoints and (b) did not receive an autism diagnosis. Participant demographics are presented in Table 1.

**Table 1.**
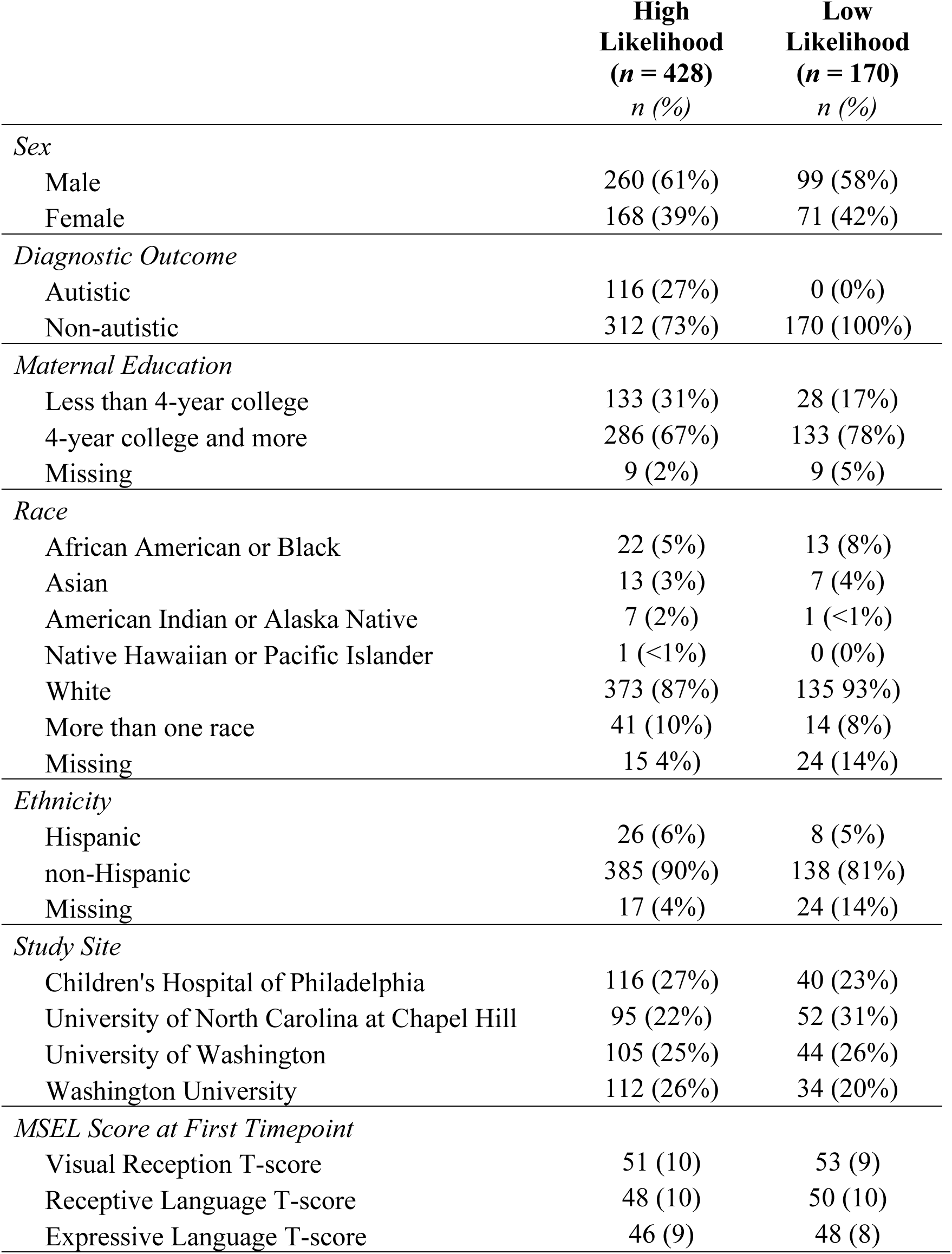
Demographics of Data Source by Familial Likelihood of Autism.

### Diagnostic Evaluations

Clinical best-estimate diagnoses were completed by licensed clinicians for all participants included in the IBIS study based on DSM-IV-TR criteria because DSM-5 diagnostic criteria were not available at the start of data collection. All available assessment data were used to formulate diagnostic outcomes at 24m, including gold-standard, autism-specific measures such as the Autism Diagnostic Observation Schedule, 2^nd^ edition (ADOS-2; Lord et al., 2012) and the Autism Diagnostic Interview-Revised (ADI-R; Rutter et al., 2003), as well as developmental information from the Mullen Scales of Early Learning (MSEL; Mullen, 1995) and the Vineland Adaptive Behavior Scales-II (VABS; Sparrow et al., 2005). ADOS-G was administered for assessments conducted before 2012, and the ADOS-2 was used for assessments after that date (Lord et al., 2000). Reliability for diagnostic decisions and across diagnostic instruments was established and maintained between sites through monthly case reviews. For a detailed protocol on diagnostic evaluations, see Estes et al. (2015).

## Measures

### Autism Diagnostic Interview-Revised (ADI-R; Rutter et al., 2003)

The ADI-R is a semi-structured, clinician-administered interview conducted with parents in the IBIS study when infants were 24- and 36-months of age. The ADI-R is designed to assess early milestones, social behavior, and repetitive behaviors/interest in support of the autism diagnostic criteria. The ADI-R also assesses the presence of regression of language or other skills. For the current study, item #11 was used to determine regression status. The ADI-R considers language regression to be a reported loss of five or more meaningful words (excluding ‘mama’ or ‘dada’). The words must have been used for at least three months and then lost for at least three months (Rutter et al., 2003). To date, this approach is the most frequent way researchers have documented early language regression (Tan et al., 2021). Of note, onset of regression was inconsistently recorded and duration beyond three months was not reported. Therefore, these factors are not considered in this paper.

### Mullen Scales of Early Learning (MSEL; Mullen, 1995)

The MSEL was used for three purposes for the current study: (1) to generate our measure-defined language regression group, (2) to assess expressive and receptive language at all time points, and (3) to complete nearest-neighbor-matching on nonverbal developmental level. The MSEL is a standardized behavioral assessment for children ages 0-68 months with subscales pertaining to expressive language, receptive language, visual reception, fine motor, and gross motor skills (Mullen, 1995). The MSEL has supporting evidence for its use as a valid and reliable measure of language in early infancy and in autistic youth (Swineford et al., 2015).

Participants met criteria for the measure-defined regression group if they had a decrease in expressive or receptive language raw scores on the MSEL between adjacent timepoints. Given a temporal gap of 3-12 months between testing sessions it is unexpected that an individual would score at or below their raw score obtained prior, which makes this approach helpful for determining a “regression”. This approach replicates previous research that used any decrease in raw scores to determine regression status (Brian et al., 2014; R. Landa, 2007; R. Landa & Garrett-Mayer, 2006; R. J. Landa et al., 2013; Ozonoff & Iosif, 2019). Additionally, T-scores, normed for corrected age in months and days, were used for both expressive and receptive language to evaluate language outcomes at 36+ months. Lastly, Visual Reception subscale T-scores are a metric of nonverbal developmental level and were used to match participants (described in more detail below).

### Sample Selection

Participants with language regression were identified within the HL group. The parent-reported language regression sample was selected based on parent endorsement of a language regression on a parent interview, the ADI-R (Rutter et al., 2003). The measure-defined regression sample was defined by a decrease in expressive or receptive language raw scores on the MSEL (Mullen, 1995) between adjacent timepoints. The average decline in scores was small at 2-3 points depending on language type investigated. Using nearest-neighbor matching, we identified one HL and one LL comparison for each regression sample (parent-reported and measure-defined), resulting in four comparison groups overall. These comparison groups were matched to the regression samples on sex, maternal education, and MSEL Visual Reception T-score at the first timepoint. Participants in the comparison samples did not have language regression or autism diagnoses.

### Participants

Twelve HL participants in the IBIS study had a parent-reported language regression on the ADI-R at 24 or 36 months. Comparison samples of non-autistic HL-match and LL-match children without language regression were chosen using nearest-neighbor matching of first timepoint Visual Reception T-score on the MSEL. Therefore, three samples were included in the aim three analyses of parent-reported language regression: regression, HL-match, and LL-match.

Thirty-six HL participants demonstrated a decrease in their receptive or expressive language raw scores between adjacent timepoints. Comparison samples of non-autistic HL-match and LL-match children without language regression were chosen using nearest-neighbor matching of first timepoint Visual Reception T-score on the MSEL. Therefore, three samples were included in the aim three analyses of measure-defined language regression: regression, HL-match, and LL-match.

One participant was present in both the parent-reported and measure-defined groups; therefore, the combined sample consists of 47 participants with language regression, 47 HL matched non-autistic, and 47 LL matched, non-autistic participants. Among these participants, age at enrollment varied between 3 and 12 months of age, and age at final visit was either 24 or 36 months of age. Visits followed staggered schedules with more frequent visits in early infancy and less frequent visits in toddlerhood. Visits could occur at three, six, nine, 12, 15, 18, 24, and 36 months. Participants in these samples completed a minimum of two visits and a maximum of five visits.

### Analytic Plan

Data preparation and analyses were conducted in R Studio (version 2024.12.0). Aim one provides descriptive statistics of the obtained regression groups and observed agreement between the groups using Cohen’s kappa. Kappa rates the observed degree of agreement, corrected for expected agreement (McHugh, 2012). The interpretation of kappa coefficients is adopted from Landis and Koch (1977): <0 = poor, .01-.20 = slight, .21-.40 = fair, .41-.60 = moderate, .61-.80 = substantial, and .81-1.00 = almost perfect.

Aim two assesses the associations between language regression and later autism diagnosis via odds ratios (ORs). Odds ratios represent the odds that an outcome will occur given a particular exposure; in this case, the odds of an autism diagnosis given the exposure of a language regression, compared to the absence of language regression (Szumilas, 2010). Odds ratios can be interpreted using the OR value and confidence intervals: OR=1 indicates exposures do not impact odds, OR>1 indicates exposures associated with higher odds, and OR<1 indicates exposures associated with lower odds. Confidence intervals at 95% are often used as a proxy to infer statistical significance if they exclude the null value (OR=1; Szumilas, 2010). To directly assess statistical significance, Fisher’s exact tests are used, as this method is robust to sparse and unbalanced cell counts. Follow-up two one-sided *t*-tests (TOST) for equivalence are used to evaluate whether the observed ORs are statistically equivalent to an OR of 1 using standard bounds of 0.5 and 2. In addition to the use of ORs to determine likelihood of autism diagnosis given a regression, we further assessed diagnostic group differences in language at the final time point using Welch’s *t*-tests, which can best account for small sample size. Language trajectories by diagnostic outcome are also visually summarized using LOESS (locally estimated scatterplot smoothing) lines of best fit (Jacoby, 2000). This approach was chosen because it provides a descriptive visualization of trends through localized regression without formal hypothesis testing or inference of statistical significance which were not possible in these small samples.

For aim 3, we examine whether developmental trajectories of raw language scores differ between the two regression groups, and their HL and LL nearest-neighbor matched samples without language regression using four sets of nested linear mixed effects models (LMM; Bates et al., 2015). LMM are appropriate for longitudinal data as they allow for modeling of both group level and individual level variability. We generate four LMM models to reflect our *two* outcomes, group differences in (1) expressive and (2) receptive language trajectories, in our *two* samples, (3) parent-reported and (4) measure-defined regression groups relative to their matched samples. We include participant level random effects to account for repeated-measures and fixed effects of age, group, and their interaction. We rescaled age for these models by subtracting the age of the youngest participant at the earliest timepoint, resulting in an age variable with a meaningful zero and interpretable model intercepts. Finally, to determine whether the presence of a language regression impacts language ability at our final timepoint, we use one-way ANOVAs to detect group differences (regression, HL-match, LL-match) in language outcomes (final timepoint T-scores) for both expressive and receptive language.

## Results

### Aim 1: Rates of Language Regression in Children at High-Likelihood for Autism

Of the high-likelihood participants in the IBIS sample with behavioral data across at least two time points (*N*=428), parent-reported regression was observed in 2.8% (*n*=12); measure-defined regression was observed in 8% (*n*=36), with only one participant having both parent-reported and measure-defined regression. Collapsing across both ways of measuring regression, 11% of high-likelihood participants (*n*=47) met criteria for language regression. Interrater agreement between parent-reported and measure-defined definitions was high by percent agreement (89%), but since this was driven by a high number of individuals without regression, Cohen’s kappa did not reveal agreement beyond chance, κ = -.0004, 95% CI [–.08, .08].

Of the measure-defined regression participants (*n*=36), 22 (61%) demonstrated a regression in receptive language, 19 (52%) demonstrated a regression in expressive language, with 5 (14%) of these demonstrating regression in both expressive and receptive language. The average measure-defined regression was a loss of 2.3 points (range=1-6) for expressive language and 3.2 points (range=1-11) for receptive language.

### Aim 2: Associations between Language Regressions and Diagnostic Outcomes

Table 2 includes a demographic breakdown of the likelihood status and autism status of the participants comprising each regression group. Participants with parent-reported regression were 2.77 times more likely to receive an autism diagnosis, compared to those without a parent-reported regression, 95% CI [0.83, 9.3], *p* = .09, and participants with measure-defined regression were 1.21 times more likely to receive an autism diagnosis compared to those who did not regress, 95% CI [0.51, 2.5], *p* = .69. Although the two odds ratios were not statistically significant as defined by an *a priori* alpha cutoff of .05, the equivalence tests also suggested non-equivalence. The 95% CI for the parent-reported and measure-defined groups extended beyond the equivalence region, 0.5 to 2, which does not support equivalence for odds ratios in parent-reported, lower *z*=2.92, *p*=.99, higher *z*=.56, *p*=.28, or measure-defined groups, lower *z*=2.31, *p*=.99, higher *z*=-1.34, *p*=.91. The presence of language regression was not associated with a significantly higher odds ratio of autism diagnosis, but the difference in this odds ratio also failed to reach statistical equivalence, indicating that this sample may not be powered to detect the relationship between language regression and diagnostic outcome.

**Table 2.**
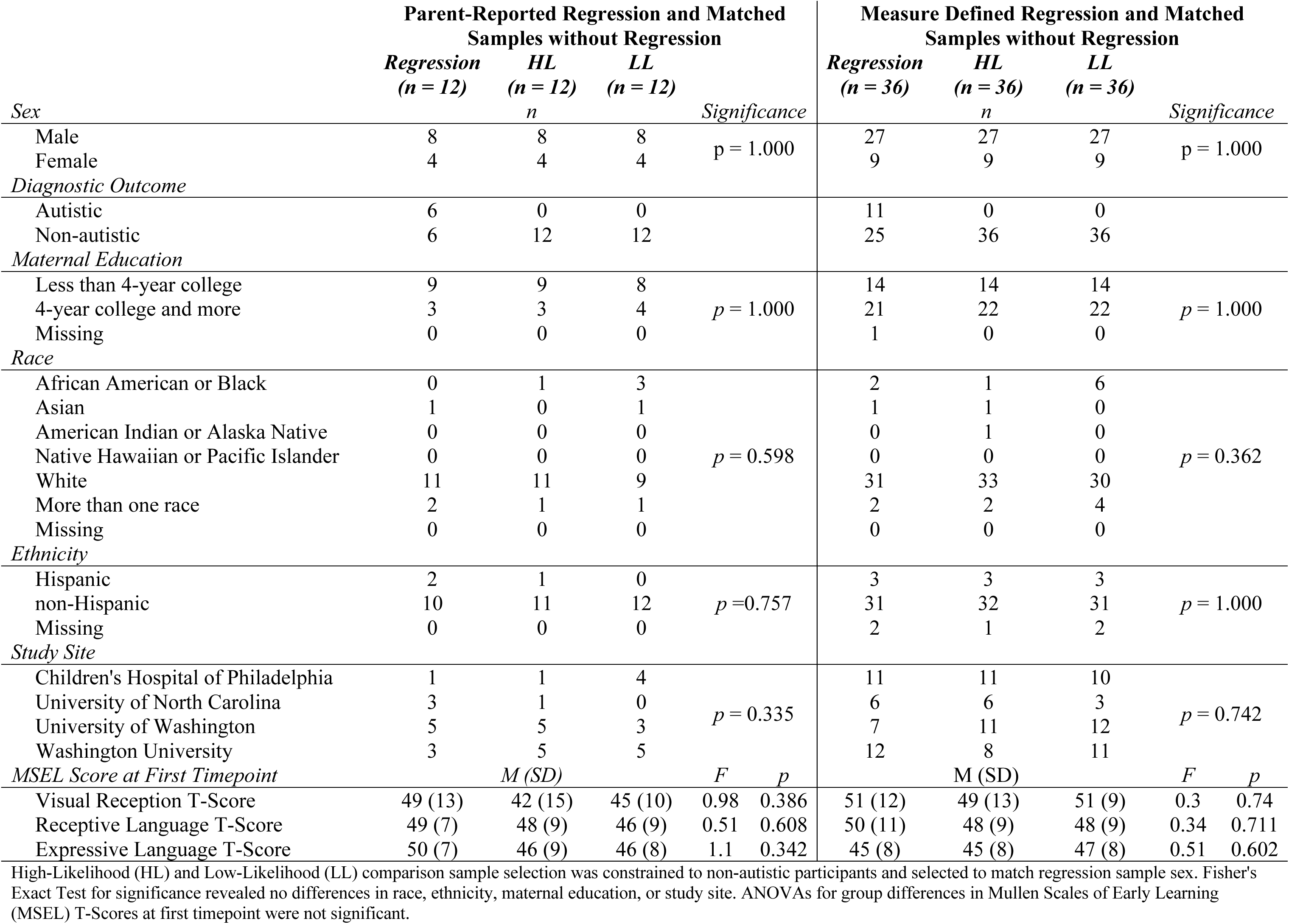
Parent-Reported and Measure-Defined Regression Groups with Matched Samples.

Figure 1 visually summarizes trajectories of expressive and receptive language raw scores by diagnostic group with LOESS lines (locally estimated scatterplot smoothing). The parent-reported regression group’s final timepoint for expressive language T-scores, *t*(9.41) = 3.55, *p* = .005, differed significantly between autistic and non-autistic participants, whereas receptive language T-scores did not, *t*(8.92) = 2.14, *p* = .06. The measure-defined regression group’s final timepoint for expressive language T-scores, *t*(10.88) = 1.90, *p* = .08, did not differ significantly between autistic and non-autistic participants, whereas receptive language T-scores did significantly differ, *t*(8.39) = 3.25, *p* = .01. Thus, among toddlers with language regression, those diagnosed with autism demonstrated some lower language scores that those not diagnosed with autism.

**Figure 1.**
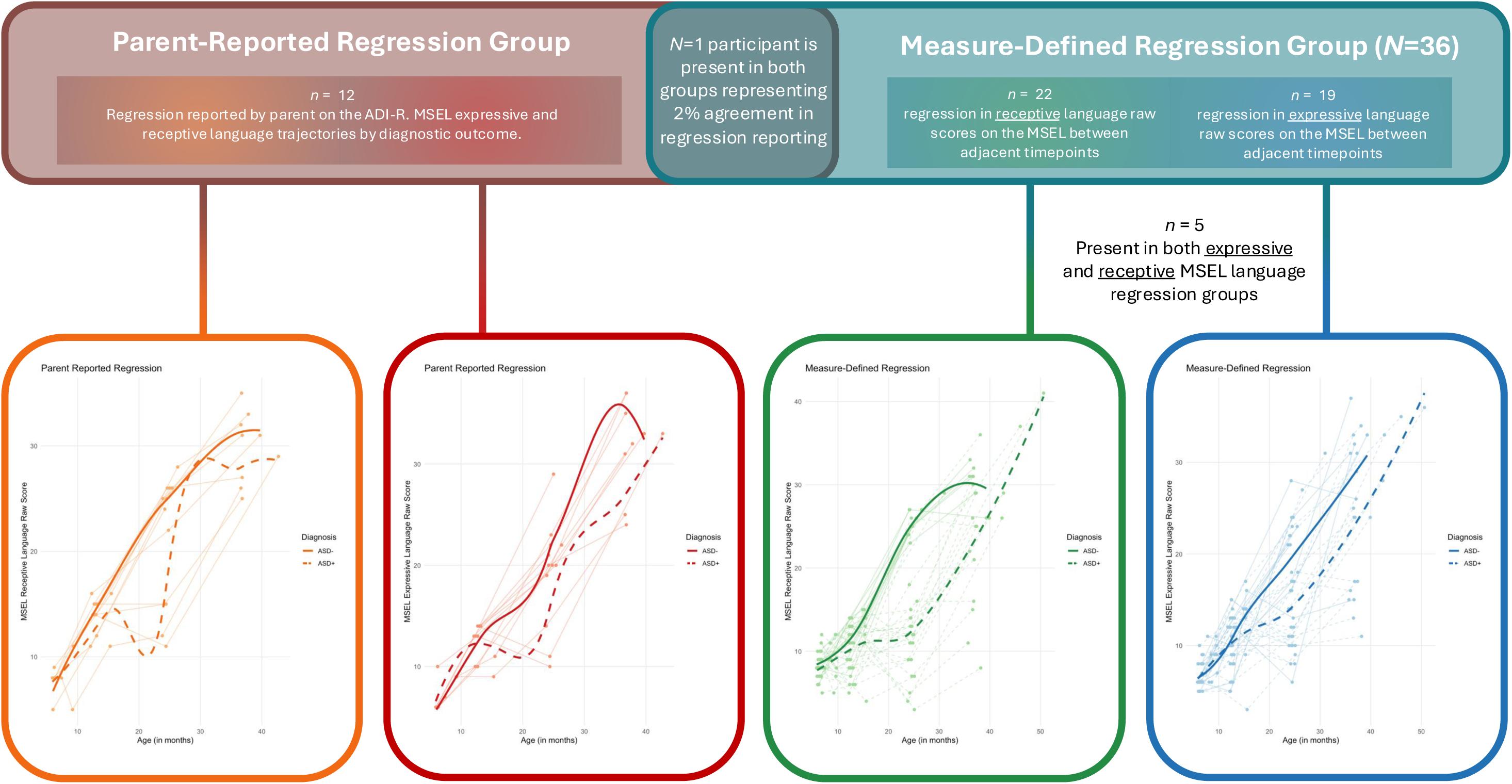
Parent-Reported and Measure-Defined Regression Groups by Diagnostic Outcome.

### Aim 3: Language Trajectories in Regression and Matched Samples

We fit four sets of 3-step nested linear mixed effects models to examine how (1) age, (2) age and group (LL-match, HL-match, Regression), and (3) the interaction of age and group predicted expressive and receptive raw score growth in the parent-reported and measure-defined regression samples compared to HL-match and LL-match samples. The language regression group was the reference group in all models. All models included a random intercept for participant to account for repeated measures. Across all models, outcomes, and samples, age was a significant positive predictor of language scores, *p*<.001, indicating growth with age.

The parent-reported regression samples evidenced a group difference in expressive language such that the LL-match group had higher expressive language scores than the regression group, *β*=2.01, *p*=.008, but this group difference was no longer significant when interactions between group and age were introduced in step three, *β*=0.32, *p*=.779. Age did not interact with group to predict different rates of expressive language development for the LL-match, *β*=0.11, *p*=.052, or HL-match groups, *β*=0.05, *p*=.347, compared to the regression group.

The parent-reported regression samples evidenced a similar group difference in receptive language such that the LL-match group had higher receptive language scores than the regression group, *β*=2.39, *p*=.008. This group difference was no longer significant in the final model because the interaction between age and group better captured between-group variance. The HL-match, *β*=0.11, *p*=.047, and LL-match groups, *β*=0.21, *p*<.001, evidenced faster receptive language growth than the regression group.

The measure-defined regression sample evidenced group differences in expressive language such that both HL-match, *β*=3.94, *p*<.001, and LL-match groups, *β*=5.24, *p*<.001, had higher expressive language scores than the regression group. Interestingly, in the final model group differences persist such that the HL-match group, *β*=-2.56, *p*=.039, had lower expressive language scores than the regression group at the earliest timepoints, but significant interactions between age and group for the HL-match, *β*=0.41, *p*<.001, and LL-match groups, *β*=0.43, *p*<.001, indicate faster expressive language growth than the regression group.

The measure-defined regression group also evidenced group differences in receptive language in the second model, such that both HL-match, *β*=5.38, *p*<.001, and LL-match groups, *β*=5.79, *p*<.001, had higher receptive language scores than the regression group. However, these group differences were no longer significant in the final model because the interactions between age and group better captured between-group variance. Age interacted with group to predict faster receptive language growth in the HL-match, *β*=0.36, *p*<.001, and LL-match groups, *β*=0.42, *p*<.001, as compared to the regression group. Full model estimates and variance components are presented in Tables 3 and 4 and LMM are presented in Figures 2 and 3.

**Figure 2.**
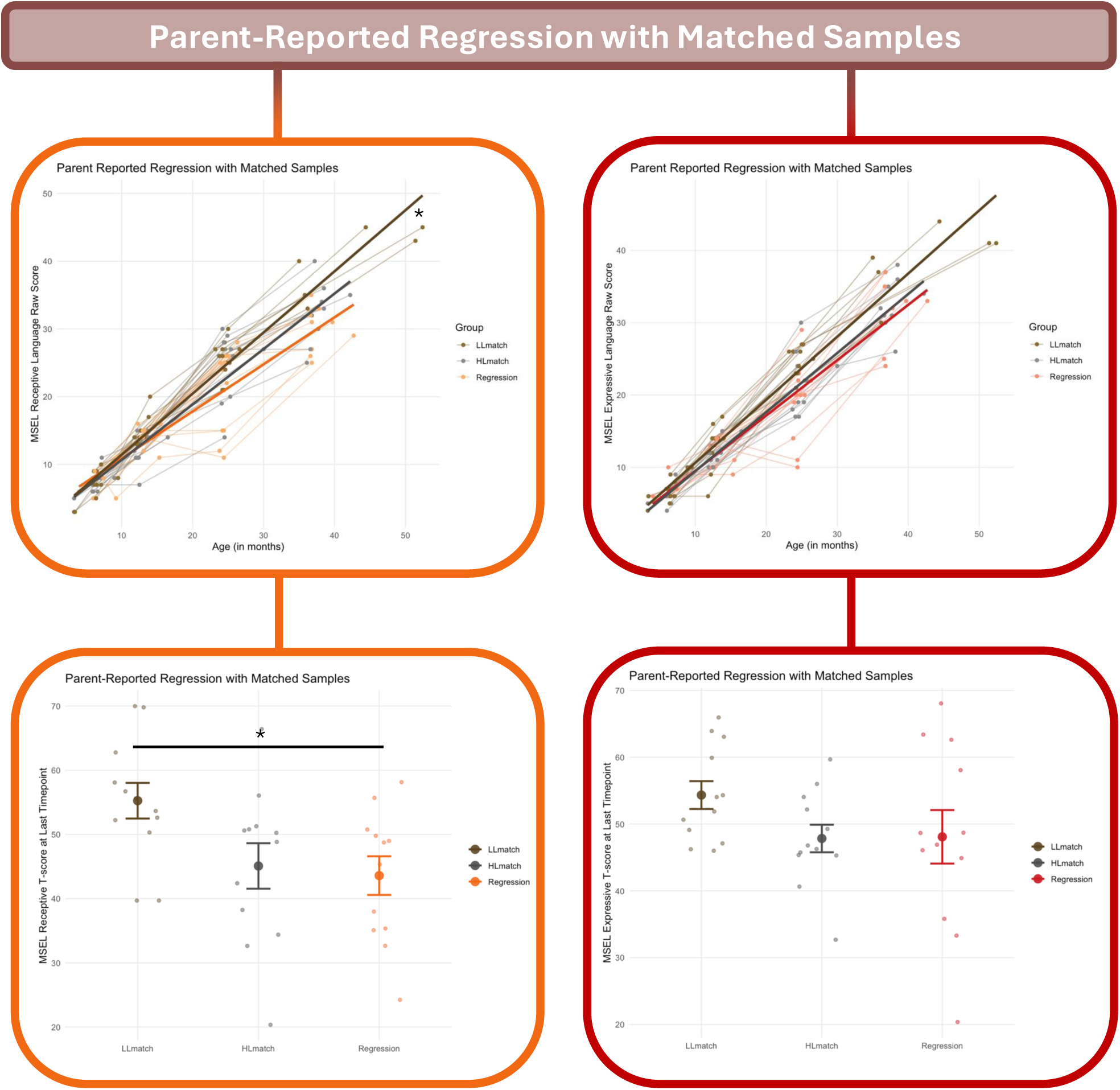
Group Differences in Language Trajectories and Outcomes for Parent-Reported Regression Group and Matched Samples.

**Figure 3.**
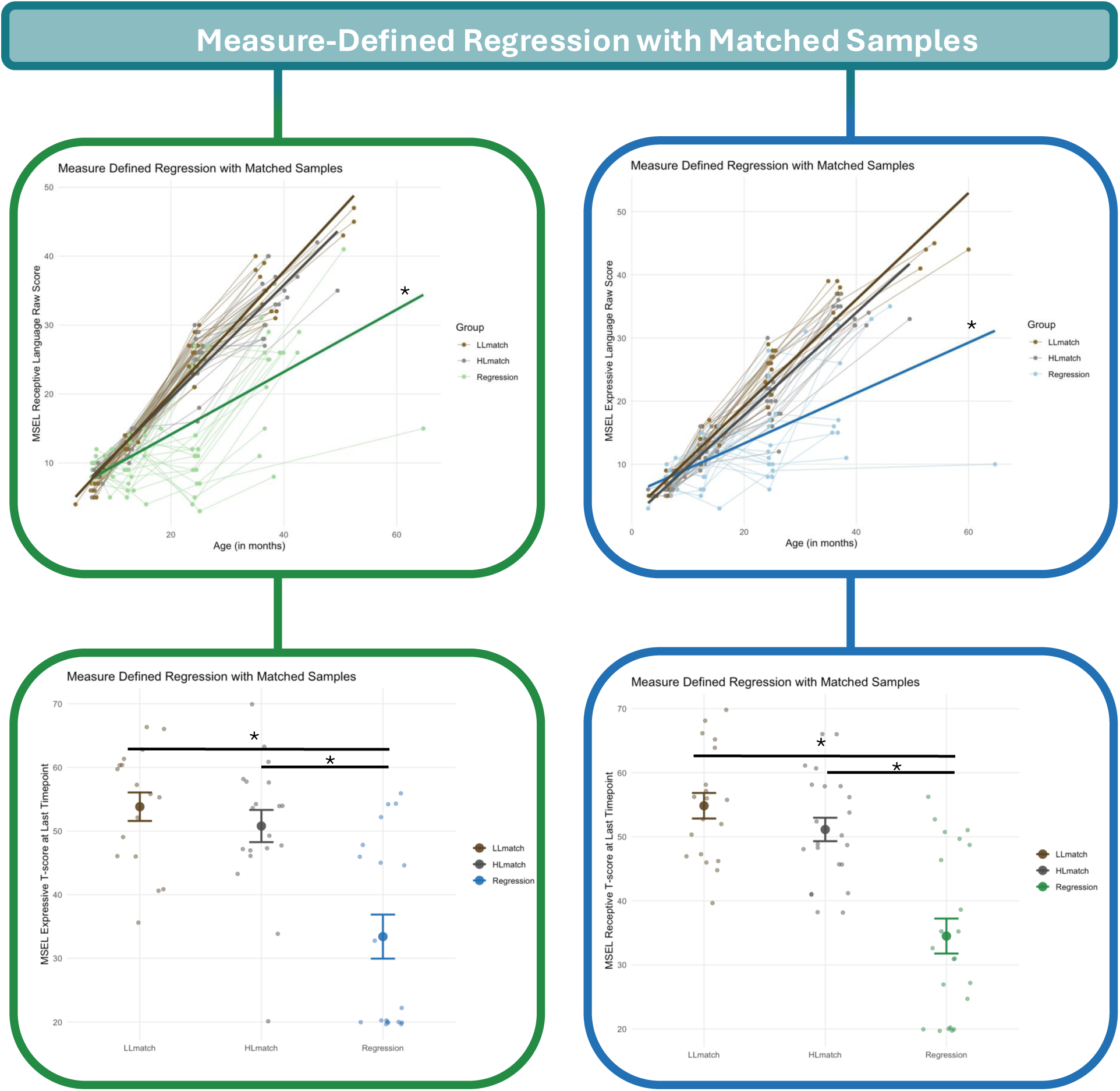
Group Differences in Language Trajectories and Outcomes for Measure-Defined Regression Group and Matched Samples.

**Table 3.** Multilevel Linear Mixed Effects Model Results for Parent-Reported Regression: Language Ability Predicted by Age, Group, and their Interaction.

| Parent-Reported Regression and Matched Groups |  |  |  |  |  |  |  |  |  |
| --- | --- | --- | --- | --- | --- | --- | --- | --- | --- |
| Predictors | Expressive Language |  |  | Expressive Language |  |  | Expressive Language |  |  |
| | $\beta$ | t-stat | p | $\beta$ | t-stat | p | $\beta$ | t-stat | p |
| Intercept | 4.37 | 9.15 | <0.001 | 3.46 | 5.29 | <0.001 | 4.45 | 5.06 | <0.001 |
| Age | 0.82 | 37.28 | <0.001 | 0.82 | 37.37 | <0.001 | 0.76 | 18.73 | <0.001 |
| Group (reference: Language Regression) |  |  |  |  |  |  |  |  |  |
| HL-match |  |  |  | 0.46 | 0.62 | 0.538 | -0.44 | -0.37 | 0.709 |
| LL-match |  |  |  | 2.01 | 2.68 | 0.008 | 0.32 | 0.28 | 0.779 |
| Age x Group (reference: Language Regression) |  |  |  |  |  |  |  |  |  |
| HL-match |  |  |  |  |  |  | 0.05 | 0.94 | 0.347 |
| LL-match |  |  |  |  |  |  | 0.11 | 1.96 | 0.052 |
| Random Effects |  |  |  |  |  |  |  |  |  |
| $\sigma^2$ | | 8.7 | | | 8.8 | | | 8.65 | |
| ICC |  | 0.15 |  |  | 0.09 |  |  | 0.09 |  |
| N |  | 36<sub>id</sub> |  |  | 36<sub>id</sub> |  |  | 36<sub>id</sub> |  |
| Observations |  | 132 |  |  | 132 |  |  | 132 |  |
| Marginal R<sup>2</sup> |  | 0.903 |  |  | 0.908 |  |  | 0.91 |  |
| Conditional R<sup>2</sup> |  | 0.918 |  |  | 0.917 |  |  | 0.918 |  |
| Predictors | Receptive Language |  |  | Receptive Language |  |  | Receptive Language |  |  |
| | $\beta$ | t-stat | p | $\beta$ | t-stat | p | $\beta$ | t-stat | p |
| Intercept | 5.52 | 10.35 | <0.001 | 4.29 | 5.73 | <0.001 | 6.27 | 6.62 | <0.001 |
| Age | 0.81 | 34.42 | <0.001 | 0.81 | 34.60 | <0.001 | 0.70 | 16.67 | <0.001 |
| Group (reference: Language Regression) |  |  |  |  |  |  |  |  |  |
| HL-match |  |  |  | 1.02 | 1.15 | 0.253 | -0.93 | -0.73 | 0.464 |
| LL-match |  |  |  | 2.39 | 2.70 | 0.008 | -0.88 | -0.71 | 0.481 |
| Age x Group (reference: Language Regression) |  |  |  |  |  |  |  |  |  |
| HL-match |  |  |  |  |  |  | 0.11 | 2.01 | 0.047 |
| LL-match |  |  |  |  |  |  | 0.21 | 3.70 | <0.001 |
| Random Effects |  |  |  |  |  |  |  |  |  |
| $\sigma^2$ | | 9.80 | | | 9.85 | | | 8.87 | |
| ICC |  | 0.22 |  |  | 0.16 |  |  | 0.19 |  |
| N |  | 36<sub>id</sub> |  |  | 36<sub>id</sub> |  |  | 36<sub>id</sub> |  |
| Observations |  | 132 |  |  | 132 |  |  | 132 |  |
| Marginal R<sup>2</sup> |  | 0.882 |  |  | 0.888 |  |  | 0.896 |  |
| Conditional R<sup>2</sup> |  | 0.907 |  |  | 0.906 |  |  | 0.916 |  |

**Table 4.** Multilevel Linear Mixed Effects Model Results for Measure-Defined Regression: Language Ability Predicted by Age, Group, and their Interaction.

| Measure-Defined Regression and Matched Groups |  |  |  |  |  |  |  |  |  |
| --- | --- | --- | --- | --- | --- | --- | --- | --- | --- |
| *Predictors* | *Expressive Language* |  |  | *Expressive Language* |  |  | *Expressive Language* |  |  |
| | $\beta$ | *t*-stat | *p* | $\beta$ | *t*-stat | *p* | $\beta$ | *t*-stat | *p* |
| *Intercept* | 4.73 | 7.30 | <0.001 | 1.78 | 2.25 | 0.025 | 6.29 | 7.17 | <0.001 |
| *Age* | 0.70 | 25.78 | <0.001 | 0.70 | 26.00 | <0.001 | 0.41 | 10.28 | <0.001 |
| *Group (reference: Language Regression)* |  |  |  |  |  |  |  |  |  |
| *HL-match* |  |  |  | 3.94 | 4.20 | <0.001 | -2.56 | -2.08 | 0.039 |
| *LL-match* |  |  |  | 5.24 | 5.41 | <0.001 | -1.53 | -1.24 | 0.217 |
| *Age x Group (reference: Language Regression)* |  |  |  |  |  |  |  |  |  |
| *HL-match* |  |  |  |  |  |  | 0.41 | 7.28 | <0.001 |
| *LL-match* |  |  |  |  |  |  | 0.43 | 7.75 | <0.001 |
| **Random Effects** |  |  |  |  |  |  |  |  |  |
| $\sigma^2$ | | 22.47 | | | 22.52 | | | 16.01 | |
| ICC |  | 0.24 |  |  | 0.09 |  |  | 0.14 |  |
| N |  | 55<sub>id</sub> |  |  | 55<sub>id</sub> |  |  | 55<sub>id</sub> |  |
| Observations |  | 207 |  |  | 207 |  |  | 207 |  |
| Marginal R<sup>2</sup> |  | 0.722 |  |  | 0.764 |  |  | 0.824 |  |
| Conditional R<sup>2</sup> |  | 0.788 |  |  | 0.786 |  |  | 0.848 |  |
| *Predictors* | *Receptive Language* |  |  | *Receptive Language* |  |  | *Receptive Language* |  |  |
| | $\beta$ | *t*-stat | *p* | $\beta$ | *t*-stat | *p* | $\beta$ | *t*-stat | *p* |
| *Intercept* | 5.60 | 8.69 | <0.001 | 2.10 | 2.86 | 0.005 | 6.25 | 7.63 | <0.001 |
| *Age* | 0.71 | 28.69 | <0.001 | 0.71 | 28.84 | <0.001 | 0.46 | 13.22 | <0.001 |
| *Group (reference: Language Regression)* |  |  |  |  |  |  |  |  |  |
| *HL-match* |  |  |  | 5.38 | 6.23 | <0.001 | -0.68 | -0.58 | 0.563 |
| *LL-match* |  |  |  | 5.79 | 6.43 | <0.001 | -1.19 | -1.00 | 0.320 |
| *Age x Group (reference: Language Regression)* |  |  |  |  |  |  |  |  |  |
| *HL-match* |  |  |  |  |  |  | 0.36 | 7.08 | <0.001 |
| *LL-match* |  |  |  |  |  |  | 0.42 | 8.08 | <0.001 |
| **Random Effects** |  |  |  |  |  |  |  |  |  |
| $\sigma^2$ | | 22.65 | | | 22.55 | | | 16.38 | |
| ICC |  | 0.29 |  |  | 0.10 |  |  | 0.15 |  |
| N |  | 63<sub>id</sub> |  |  | 63<sub>id</sub> |  |  | 63<sub>id</sub> |  |
| Observations |  | 243 |  |  | 243 |  |  | 243 |  |
| Marginal R<sup>2</sup> |  | 0.714 |  |  | 0.775 |  |  | 0.826 |  |
| Conditional R<sup>2</sup> |  | 0.797 |  |  | 0.796 |  |  | 0.852 |  |

Expressive language T-scores at the final timepoint did not differ between parent-reported regression and matched samples, *F*(2,33)=1.65, *p*=.208. Receptive language T-scores in the parent-reported regression group were significantly different, *F*(2,33)=4.12, *p*=.025. Interestingly, post-hoc comparisons using Tukey’s HSD indicating that the LL-match group had significantly higher receptive language than the regression group, *p*=.03, but the regression group, *p*=.94, and LL-match group, *p*=.07, did not differ from the HL-match group.

Expressive language T-scores significantly differed between the measure-defined regression group and HL-match and LL-match groups, *F*(2,52)=15.24, *p*<.001. Post-hoc comparisons indicated that children in the regression group had significantly lower expressive language T-scores than both the LL-match group, *p*<.001, and the HL-match group, *p*<.001. There was no significant difference between HL-match and LL-match, *p*=.74. Similarly, receptive language T-scores also showed significant group differences, *F*(2,60)=23.43, *p*<.001. Post-hoc comparisons indicated that the regression group had significantly lower receptive language scores than both the LL-match group *p*<.001 and the HL-match group *p*<.001, with no significant difference between HL-match and LL-match, *p*=.49. Boxplots showing these differences in final timepoint receptive and expressive language T-scores are presented in Figure 2 for the parent-reported ANOVA and Figure 3 for measure-defined ANOVA.

## Discussion

The current study sought to characterize language regressions using parent-reported and measure-defined constructs in early infancy and toddlerhood. In this longitudinal sample, at high familial likelihood of autism, 11% of toddlers met criteria for language regression, combined across parent-report and measure-defined indicators. This was expectedly lower than 20-40% prevalence estimates in diagnosed autistic samples (Barger et al., 2013; Tan et al., 2021). While this prevalence rate does not reflect the presence of language regression in community ascertained samples, it may reflect the prevalence of language regression in high-likelihood samples. Having a language regression did not significantly increase the child’s odds of receiving a later autism diagnosis; however, for toddlers who did receive an autism diagnosis, the presence of a language regression contributed to significantly lower expressive and receptive language scores at 36+ months. Toddlers with parent-reported and measure-defined regression showed different language growth trajectories relative to nearest-neighbor matched groups without language regression, but all showed continued language growth and recovery of skills after regression. Interestingly, toddlers with parent-reported regression often were indistinguishable from their high-likelihood-group-matched peers by 36+ months. In contrast, toddlers with a measure-defined regression emerged as phenotypically distinct from their high-likelihood-group-matched peers on both expressive and receptive language at 36+ months. Each of these results are discussed in greater detail below.

Surprisingly, the concordance between parent-report and measure-defined groups was low. This may suggest that parents and measures are detecting two different events in language development. Future work is needed to assess alignment between other language assessment tools and parent-report and potentially develop an objective measure of language regression as proposed by Tan et al. (2021). This low concordance could also support Clarke’s theory that parents are sensitive to stagnate growth rather than *measurable loss:* “individuals with autism spectrum disorder are not ‘losing’ words, but instead are not progressing in their ability to learn and use new words” (Clarke, 2019, p. 1). These results could suggest that the iterative and cyclical nature of skill acquisition in non-autistic development (Corbetta et al., 2018) might sometimes be experienced by caregivers as skill loss. A familiar example of this blurry boundary between iterative learning and regression is toilet training. It is common for caregivers to anticipate and expect “regressions” or episodes of returning to a baseline in the iterative process of gaining some skills. Additionally, expressive language is more salient than receptive and so the wording on the ADI-R and similar tools may primarily detect expressive rather than receptive slowing or loss in skill.

Interestingly, neither parent-reported nor measure-defined regression indicators were associated with greater odds of an autism diagnosis; however, even with this large sample, the study may not have been adequately powered to identify significant differences. While our results suggest that high likelihood toddlers with and without subsequent autism diagnoses exhibit language regression, autism diagnosis was associated with lower language ability at 36+ months across both parent-reported and measure-defined regression. These different language outcomes by diagnostic group indicate that the complex impact of autism on language development extends beyond the observable symptom of language regression and that presence of language regression alone is a weak predictor of later development.

We draw the conclusion that these two forms of regression data, parent-reported and measure-defined, may capture unique events because the two types of regression were associated with different language trajectories relative to toddlers without regression. The parent-reported groups eventually “caught up” in the current study, defined by being statistically indistinguishable from their likelihood-group-matched peers, matching results by Pickles et al. (2022), but the measure-defined group demonstrated a significant phenotypic difference from their likelihood-group-matched peers. This may indicate that these two definitions of regression represent distinct patterns of language development associated with different outcomes. Furthermore, this may signal that toddlers with measure-defined regressions, or drops in raw scores across timepoints, could benefit from monitoring. Our measure-defined approach may have identified some “late talkers” explaining why language skills might still be delayed relative to peers at age three (Fisher, 2017). Importantly, it is possible that the language trajectories in the measure-defined group reflect poor tolerance of testing and more variable performance overall rather than a true loss of skills as the outcome measure of language development was also the measure used to define language regression in this group. Future research can support or refute this possibility by examining language ability across multiple types of assessments.

One important finding to note is that across all levels of all linear mixed effects models, age positively predicted language development. Toddlers with language regression in these samples kept growing in both receptive and expressive language. This finding aligns with previous work on language outcomes after regression (Goldberg et al., 2003; Pickles et al., 2009; Prescott & Weismer, 2022) and demonstrates that a regression does not mean a halt in development or permanently stalled language growth. This finding may seem intuitive for clinicians and child development researchers, but to parents experiencing alarm at their child’s skill loss, this fact could be extraordinarily reassuring. Children with language regression are still developing, growing, and working towards language mastery.

### Limitations

This study has numerous strengths, including a large, well-characterized sample, prospective design with multiple time-points between the ages of 6- and 36-months, use of well-matched comparison groups, and multi-informant modalities (e.g., parent-report, clinician report, and objective measures). Considering these strengths, a few limitations are also noted. First, while regression has previously been defined in research as a 1-point or greater drop on a standardized language assessment (Brian et al., 2014), this variation in testing performance could be a noisy indicator of regression. Infant performance on an assessment can vary due to external factors like time of day and internal factors such as attention, motivation, or illness. Additionally, the test-retest reliability for the MSEL specifically is unknown (Mullen, 1995).

Another limitation is the timing of observed and reported language regression. Parent-reported regression on the ADI-R is defined as a loss of five words that were previously used for three or more months. Parents reported regressions on the ADI-R as early as 13 months and decreases in MSEL scores occurred as early as six months. While these earliest examples of evidence of skill loss are not reflective of the timing within the majority of this sample, it is clear that both of these indicators of regression are capturing a broader loss of language skill that may occur prior to a child ability to produce five words consistently for several months (Goldberg et al., 2003; Pickles et al., 2009; Prescott & Weismer, 2022).

Another limitation of these data is the high variability in number of timepoints and spread of those timepoints. The IBIS project was designed to recruit participants on staggered schedules to enable continuous modeling of growth. The variability in number of timepoints per infant was, in part, a result of study design. Another source of variability is a wide age range for the final timepoint in the study. We refer to this timepoint as 36+ months, but due to scheduling challenges the oldest child completing the MSEL in this sample was just over five years old. Growth trajectories are modeled on raw scores and ages while final timepoint comparisons are made on T-scores due to this wide window.

Although the longitudinal direct observation of language skills concurrent with parent report in this study is illuminating, the lack of follow-up data through school age is a limitation. Previous work has encouragingly found school-age outcomes of children with and without regression to converge beyond the age range of our sample (Goldberg et al., 2003; Pickles et al., 2009; Prescott & Weismer, 2022). Future work from this research group will include following these youth through school age to evaluate language growth.

### Conclusion

Language regression occurs in infants and toddlers at elevated familial likelihood for autism at lower rates than in studies of autism diagnosed samples. Parent-reported and measure-defined regression represent distinct developmental events that rarely co-occur. The presence of language regression may not be predictive of later autism diagnosis. When compared to peers, parent-reported regression is associated with less delayed language development than measure-defined language regression.

## Data Availability

Data is not available.

## Acknowledgements

The Infant Brain Imaging Study (IBIS) Network is an NIH funded Autism Centers of Excellence project. The IBIS 1 and 2 studies consisted of a consortium of 9 universities in the U.S. and Canada. Clinical Sites: University of North Carolina: J. Piven (IBIS Network PI), H. C. Hazlett, C. Chappell; University of Washington: S. Dager, A. Estes, D. Shaw; Washington University: K. Botteron, R. McKinstry, J. Constantino, J. Pruett: Children’s Hospital of Philadelphia: R. Schultz, J. Pandey: University of Alberta: L. Zwaigenbaum: University of Minnesota; J. T. Elison, J. Wolff. Data Coordinating Center: Montreal Neurological Institute: A. C. Evans, D. L. Collins, G. B. Pike, V. Fonov, P. Kostopoulos, S. Das, L. MacIntyre. Image Processing Core: New York University: G. Gerig: University of North Carolina: M. Styner. Statistical Analysis Core: University of North Carolina: H. Gu.

